# Non-Pharmaceutical Interventions and COVID-19 Burden in the United States

**DOI:** 10.1101/2021.09.26.21264142

**Authors:** Michael J. Ahlers, Hilary J. Aralis, Wilson L. Tang, Jeremy B. Sussman, Gregg C. Fonarow, Boback Ziaeian

## Abstract

**Background:** Non-pharmaceutical interventions (NPIs) are mitigation strategies used to reduce the spread of transmissible diseases. The relative effectiveness of specific NPIs remains uncertain.

**Methods:** We used state-level Coronavirus disease 2019 (COVID-19) case and mortality data between January 19, 2020 and March 7, 2021 to model NPI policy effectiveness. Empirically derived breakpoints in case and mortality velocities were used to identify periods of stable, decreasing, or increasing COVID-19 burden. The associations between NPI adoption and subsequent decreases in case or death velocities were estimated using generalized linear models accounting for weekly variability shared across states. State-level NPI policies included: stay at home order, indoor public gathering ban (mild >10 or severe ≤10), indoor restaurant dining ban, and public mask mandate.

**Results:** 28,602,830 cases and 511,899 deaths were recorded. The odds of a decrease in COVID-19 case velocity were significantly elevated for stay at home (OR 2.02, 95% CI 1.63-2.52), indoor dining ban (OR 1.62, 95% CI 1.25-2.10), public mask mandate (OR 2.18, 95% CI 1.47-3.23), and severe gathering ban (OR 1.68, 95% CI 1.31-2.16). In mutually adjusted models, odds remained elevated for stay at home (AOR 1.47, 95% CI 1.04-2.07) and public mask mandate (AOR = 2.27, 95% CI 1.51-3.41). Stay at home (OR 2.00, 95% CI 1.53-2.62; AOR 1.89, 95% CI 1.25-2.87) was also associated with greater likelihood of decrease in death velocity in unadjusted and adjusted models.

**Conclusions:** NPIs employed in the U.S. during the COVID-19 pandemic, most significantly stay at home orders, were associated with decreased COVID-19 burden.

## Introduction

Non-pharmaceutical interventions (NPIs) are mitigation strategies that have been used to control the spread of transmissible diseases, epidemics, and pandemics for more than one hundred years.^1,2^ During the Coronavirus disease 2019 (COVID-19) pandemic, a variety of NPIs were adopted and discontinued by state governments. Strategies in the U.S. included travel bans, declarations of emergency situations, social distancing campaigns, self-quarantine upon infection or known exposure, and universal facial masking recommendations.^3,4^ These strategies were adopted and discontinued with varying timing and relation to the severity of the pandemic across U.S. states. Despite efforts to mitigate COVID-19 spread, the U.S. has reported more than 34 million cases and 600,000 deaths related to COVID-19.^5^ In 2020, COVID-19 was the third leading cause of death in the U.S. and reduced life expectancy by one year.^6^

The effectiveness of NPIs on respiratory illnesses is unclear.^7–12^ Most studies have examined NPIs individually, not accounting for multiple interventions over time.^13–15^ The purpose of this study was to evaluate the adoption and discontinuation of four broadly used NPIs (stay at home order, indoor restaurant dining ban, public mask mandate, and indoor public gathering ban) on shifts in the COVID-19 burden among U.S. states.

## Methods

### Study design, setting, and population

We performed a retrospective, observational cohort study of the U.S. population between January 19, 2020 (first diagnosed U.S. case) and March 7, 2021. The primary analysis evaluated the dates of state-specific NPI adoption or discontinuation on the COVID-19 case and mortality velocities.

### Data Collection

State-level case and mortality counts per day were obtained from The COVID Tracking Project^16^, which aggregated public reports of COVID-19 diagnostic and death certification data for the U.S. The data reported is obtained from state public health authorities or official media accounts.^13,17^

Dates for adoption and discontinuation of NPIs studied were obtained from publicly available reports (**Supplemental Table S1**).^13,18–23^ Policies that were incorporated into the analysis included stay at home order, indoor public gathering ban (mild or severe), indoor restaurant dining ban, and public mask mandate. Dates for each policy adoption or discontinuation were recorded from February 21, 2020 to January 29, 2021 for the time-dependent modeling. When sources were not in agreement on the date of adoption or discontinuation, executive orders and media outlet publications were reviewed.

Definitions were set for each NPI used. Indoor public gathering bans with a maximum of 10 or fewer were classified as severe, while indoor public gathering bans with maximums greater than 10 were classified as mild. Indoor restaurant dining ban was defined as adopted when indoor dining was banned and discontinued when indoor dining was reinstated, regardless of capacity specification or outdoor dining policies. Public mask mandate also applied to indoor mandates, regardless of outdoor policy. Stay at home order was defined as any statewide policy ordering discontinuation of all nonessential travel from home.

### Statistical Analysis

Each state’s daily case and death counts were used to model velocities. Time zero was defined at the establishment of community transmission (100 or more confirmed cases for each state). Disease burden over time was modeled using case (and death) velocity, obtained by taking the first derivative with respect to time of the log cumulative daily case (or death) counts reported at the state-level.^24^ Cubic splines were fit to the log cumulative counts to facilitate precise calculation of the first derivative with respect to time. A log link was applied to map the case (or death) velocities to the entire real line which allowed for modeling of velocities as a linear function.

A breakpoint analysis was used to evaluate changes in case velocity attributable to policy adoptions or discontinuations. This approach identifies time periods with deviations in the COVID-19 case or death velocity rates. A change in COVID-19 growth rate is determined if adding a new regression slope at a specific date decreases the residual sum of squares sufficiently to improve the Bayesian Information Criterion. Breakpoints were empirically identified for each state using the *strucchange* R package for dating structural changes in regression models.^25^ A minimal segment length of 14 days was set by specifying a minimum number of observations per segment (14). As a result of the minimal segment size specification, each state had at most one breakpoint per 7-day week. This allowed for determination of whether a state experienced an increase, decrease, or no change in each week.

Assessments of the impact of policy adoption assumed pre-specified time lags related to the natural course of transmission and disease. We estimated lags of 3 to 10 days from policy adoption to changes in transmission, 5 to 10 days from infection to case diagnosis, and 6 to 15 days from case diagnosis to death (**Supplemental Material: COVID-19 Natural History Estimations**). Based on these intervals, policies adopted between 7 to 21 days (1-3 weeks) prior to the start date in each week were considered when evaluating impacts on case velocity. Policies adopted between 14 and 35 days (2-5 weeks) prior to the start date of a given week were considered when evaluating impacts on death velocity. Generalized linear models using generalized estimating equations (GEE) were used to evaluate policy changes in relation to shifts in COVID-19 case and death velocities. A 3-level ordered outcome variable for case and death velocities (increase, no change, or decrease) was used. Ordinal logistic regression with a cumulative logit link function modeled the probability of a state-week corresponding to (1) a decrease in case (or death) velocity vs. no change and (2) no change vs. an increase in case (or death) velocity.

An independent working correlation structure was specified to account for within-state correlation. Two sets of models were fit including: (1) a fixed effect for the given policy being modeled (a separate unadjusted model for each policy) (2) a fixed effect for each policy (one adjusted model for all policies). Time variability shared across states was accounted for by including week-number as a continuous fixed effect and utilizing three change-points to allow for piecewise trends during phases of the pandemic. We aligned four phases with the calendar weeks being modeled (**Supplemental Material: Model Phasing)**.

Policy models were fit to data consisting of one observation per state, per week starting with the first week in which a breakpoint was observed. To summarize results from the cumulative logit model formulation, we use the phrase *decrease in case velocity* to imply either the probability of observing a decreasing breakpoint vs. no change or observing no change vs. an increasing breakpoint. Analyses were performed in R 4.0.3 (R Foundation for Statistical Computing, Vienna, Austria).^26^

## Results

During the period studied, there were 28,602,830 cases and 511,899 deaths recorded. A total of 409 NPI adoptions and discontinuations were recorded (**Supplemental Table S1**). We recorded and ranked states by total cases per capita as of March 7, 2021 (**Supplemental Table S2**). Timelines for the three highest and lowest case burden states are depicted as examples of NPI utilization variability (**Figure 1**).

**Figure 1.**
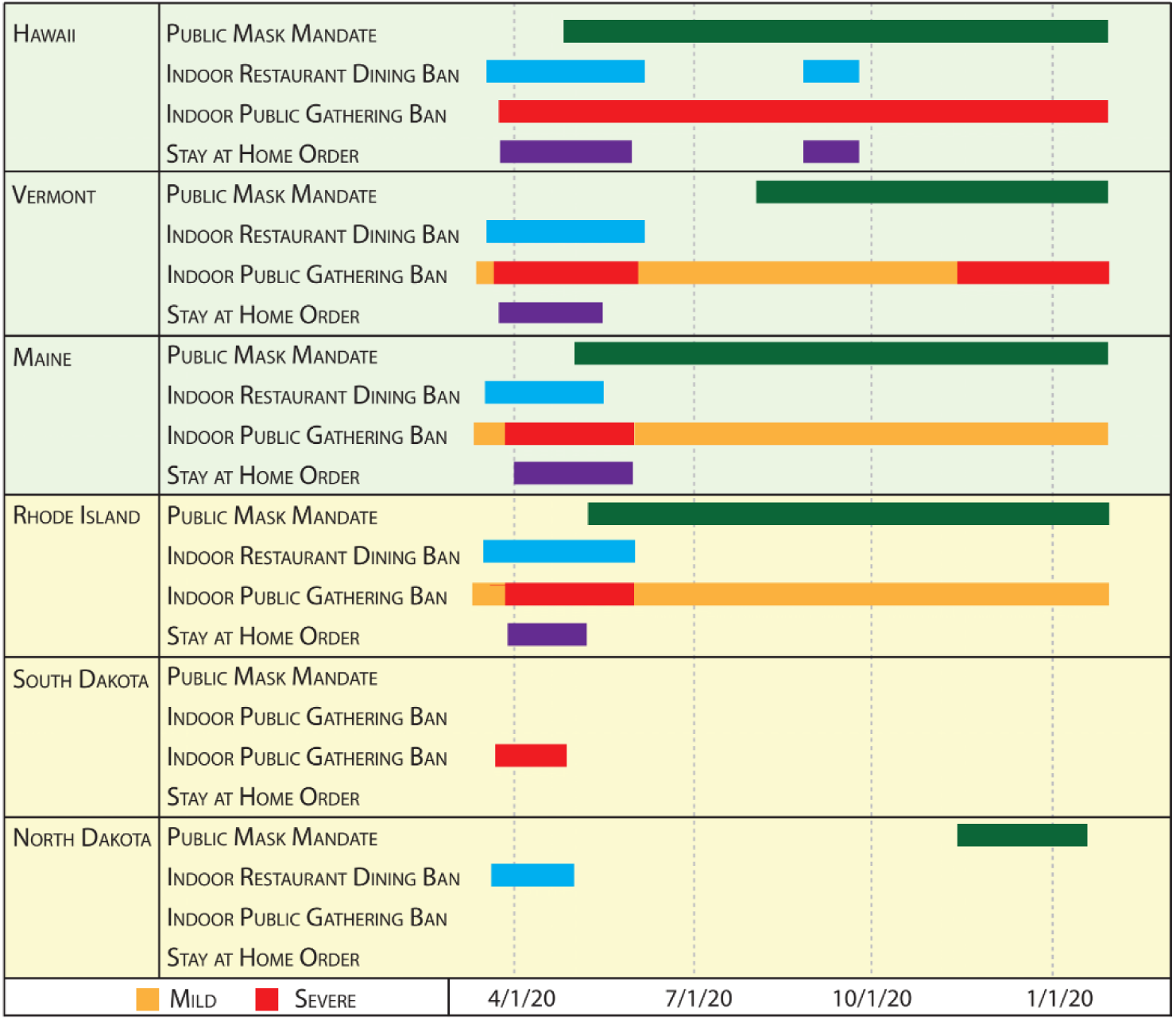
Timeline of non-pharmaceutical interventions for top 3 lowest (light green) and highest (light yellow) U.S. states by COVID-19 cases per capita. “Mild” refers to indoor public gathering bans with maximums greater than 10. “Severe” refers to indoor public gathering bans with maximums of 10 or fewer. States in a background of light green represent the top 3 states with the lowest number of COVID-19 cases per capita as of March 7, 2021 (Hawaii 2,020 per 100,000, Vermont 2,568, and Maine 3,422). States in a background of light yellow represent the top 3 states with the highest number of COVID-19 cases per capita as of March 7, 2021 (Rhode Island 12,180 per 100,000, South Dakota 12,875, and North Dakota 13,208).

### NPI influence of COVID-19 Case Velocities

A total of 603 case breakpoints were identified (**Table 1**). A decrease in case velocity was observed in 433 (71.8%) breakpoints and an increase in case velocity was observed in 170 (28.2%). Across all 50 states, the median number of case breakpoints was 12 (minimum of 7 and maximum of 17). The quantity and type of breakpoint was graphed chronologically (**Figure S2**). Cumulative cases over time were plotted along with their estimated breakpoints for the three best and worst performing states according to cases per capita (**Figure 2**). Plots of the derivative of the logarithm of cumulative cases (velocity) and the logarithm of the derivative of the logarithm (data implemented in modeling) for these states is available in the supplementary material (**Figures S4 and S5**).

**Table 1.** Non-pharmaceutical interventions status relative to case and death state-week.

| NPI Type and Status | Count of Breakpoint Types |  |  |  |  |  |
| --- | --- | --- | --- | --- | --- | --- |
|  | Case Velocity<br>(2350 State-Weeks) |  |  | Death Velocity<br>(2350 State-Weeks) |  |  |
|  | Decrease<br>(N = 433) | No Change<br>(N = 1747) | Increase<br>(N = 170) | Decrease<br>(N = 330) | No Change<br>(N = 1889) | Increase<br>(N = 131) |
|  | *n* (%) | *n* (%) | *n* (%) | *n* (%) | *n* (%) | *n* (%) |
| Stay at Home Order On | 45 (10.4) | 72 (4.1) | 0 (0.0) | 53 (16.1) | 103 (5.5) | 0 (0.0) |
| Stay at Home Order Off | 22 (5.1) | 84 (4.8) | 9 (5.3) | 33 (10.0) | 116 (6.1) | 4 (3.1) |
| Indoor Restaurant Dining Ban On | 52 (12.0) | 109 (6.2) | 1 (0.6) | 57 (17.3) | 157 (8.3) | 2 (1.5) |
| Indoor Restaurant Dining Ban Off | 32 (7.4) | 112 (6.4) | 12 (7.1) | 39 (11.8) | 161 (8.5) | 6 (4.6) |
| Public Mask Mandate On | 29 (6.7) | 79 (4.5) | 3 (1.8) | 23 (7.0) | 121 (6.4) | 4 (3.1) |
| Mild Indoor Public Gathering Ban On | 38 (8.8) | 154 (8.8) | 18 (10.6) | 59 (17.9) | 210 (11.1) | 10 (7.6) |
| Severe Indoor Public Gathering Ban On | 62 (14.3) | 123 (7.0) | 1 (0.6) | 65 (19.7) | 177 (9.4) | 4 (3.1) |
| Indoor Public Gathering Ban Off | 5 (1.2) | 35 (2.0) | 4 (2.4) | 8 (2.4) | 47 (2.5) | 2 (1.5) |

N = total number of state-weeks, *n* = number of state weeks of each breakpoint type that fell within the observation period for an NPI adoption or discontinuation (7-21 days for cases, 14-35 days for deaths) % = percentage of state-weeks. Breakpoint segments were categorized as decreasing, no change, or increasing for the case and death velocity analysis.

**Figure 2.**
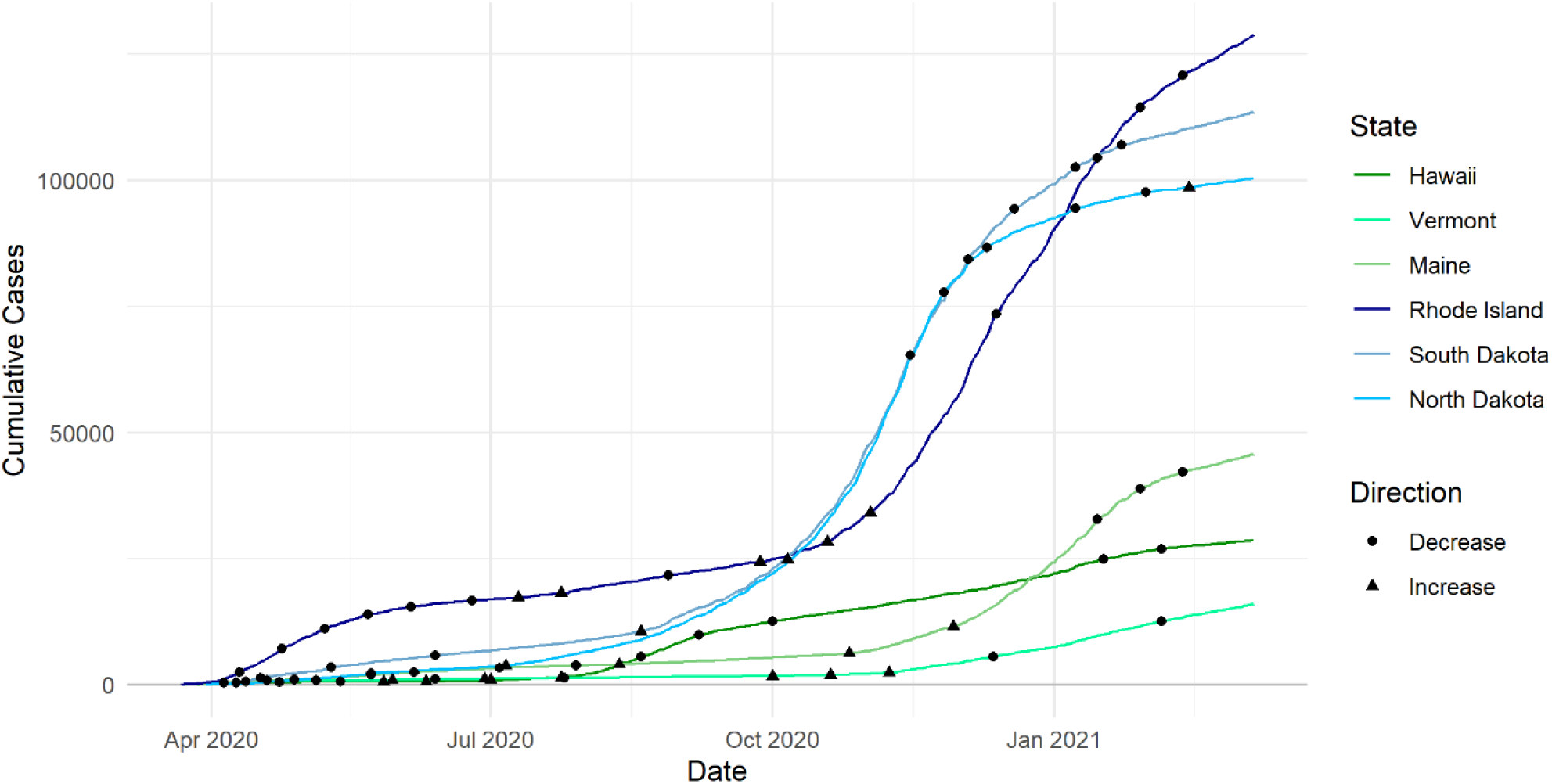
Cumulative COVID-19 cases and the breakpoints identified for the top 3 lowest and highest U.S. states by COVID-19 cases per capita. Hawaii, Vermont, and Maine ranked as the three best performing states by cumulative number of COVID-19 cases per capita as of March 7, 2021. Rhode Island, South Dakota, and North Dakota ranked as the three worst performing states over the same period. Breakpoints, dates at which the linear segments of COVID-19 case velocities showed substantial change in their rate, are plotted over the liner plot of cases for each respective state.

Ordinal logistic regression models including a single policy adoption or discontinuation estimated significantly increased odds of a decrease in case velocity following institution of stay at home orders (OR 2.02, 95% CI 1.63-2.52), indoor restaurant dining ban (OR 1.62, 95% CI 1.25-2.10), public mask mandates (OR 2.18, 95% CI 1.47-3.23), and severe indoor public gathering bans (OR 1.68, 95% CI 1.31-2.16). Institution of a mild gathering ban was associated with decreased odds of a decrease in case velocity (OR 0.51, 95% CI 0.39-0.68) (**Table 2**). In mutually adjusted models, institution of a stay at home order was associated with decreasing case velocity (adjusted odds ratio [AOR] 1.47, 95% CI 1.04-2.07), as was institution of a public masking mandate (AOR 2.27, 95% CI 1.51-3.41). Adoption of a mild indoor public gathering ban was associated with a subsequent increase in case velocities (AOR 0.46, 95% CI 0.34-0.61). (**Table 3**).

**Table 2.** Non-pharmaceutical interventions and the odds of decreasing COVID-19 case and death burden: one model per intervention.

|  | Cases |  |  | Deaths |  |  |
| --- | --- | --- | --- | --- | --- | --- |
|  | OR | 95% CI | p-value | OR | 95% CI | p-value |
| Stay at Home Order On | 2.02 | 1.63, 2.52 | **< .0001** | 2.00 | 1.53, 2.62 | **< .0001** |
| Stay at Home Order Off | 0.87 | 0.53, 1.42 | .5741 | 1.21 | 0.89, 1.63 | .2194 |
| Indoor Restaurant Dining Ban On | 1.62 | 1.25, 2.10 | **.0003** | 1.50 | 1.15, 1.95 | **.0024** |
| Indoor Restaurant Dining Ban Off | 0.94 | 0.60, 1.45 | .7652 | 1.04 | 0.78, 1.41 | .7641 |
| Public Mask Mandate On | 2.18 | 1.47, 3.23 | **.0001** | 1.39 | 0.94, 2.05 | .1015 |
| Mild Indoor Public Gathering Ban On | 0.51 | 0.39, 0.68 | **< .0001** | 0.86 | 0.62, 1.18 | .3405 |
| Severe Indoor Public Gathering Ban On | 1.68 | 1.31, 2.16 | **< .0001** | 1.45 | 1.12, 1.88 | **.0044** |
| Indoor Public Gathering Ban Off | 0.64 | 0.29, 1.40 | .2683 | 1.19 | 0.65, 2.19 | .5728 |

OR = odds ratio. CI = confidence interval. An odds ratio greater than one is associated with an increased probability that COVID-19 case or death velocities decreased. Each NPI was evaluated in isolation with an individual model. Indoor public gathering bans with a maximum of 10 or fewer were classified as severe. Indoor public gathering bans with maximums greater than 10 were classified as mild. Indoor restaurant dining ban was defined as adopted when indoor dining was banned and discontinued when indoor dining was reinstated, regardless of capacity specification or outdoor dining policies.

**Table 3.** Non-pharmaceutical interventions and the odds of decreasing COVID-19 case and death burden: one model including all interventions.

|  | Cases |  |  | Deaths |  |  |
| --- | --- | --- | --- | --- | --- | --- |
|  | Adjusted Odds Ratio |  |  | Adjusted Odds Ratio |  |  |
|  | AOR | 95% CI | p-value | AOR | 95% CI | p-value |
| Stay at Home Order On | 1.47 | 1.04, 2.07 | **.0284** | 1.89 | 1.25, 2.87 | **.0027** |
| Stay at Home Order Off | 0.93 | 0.56, 1.55 | .7911 | 1.28 | 0.89, 1.85 | .1850 |
| Indoor Restaurant Dining Ban On | 1.47 | 0.96, 2.26 | .0731 | 1.15 | 0.76, 1.74 | .5024 |
| Indoor Restaurant Dining Ban Off | 1.25 | 0.77, 2.03 | .3654 | 1.13 | 0.81, 1.59 | .4781 |
| Public Mask Mandate On | 2.27 | 1.51, 3.41 | **< .0001** | 1.45 | 0.97, 2.17 | .0670 |
| Mild Indoor Public Gathering Ban On | 0.46 | 0.34, 0.61 | **< .0001** | 0.78 | 0.56, 1.09 | .1480 |
| Severe Indoor Public Gathering Ban On | 1.38 | 0.97, 1.95 | .0738 | 1.08 | 0.72, 1.64 | .6924 |
| Indoor Public Gathering Ban Off | 0.64 | 0.30, 1.39 | .2608 | 1.16 | 0.62, 2.17 | .6426 |

AOR = adjusted odds ratio. CI = confidence interval. An adjusted odds ratio greater than 1 is associated with increased probability that COVID-19 case or death velocities decreased. All four NPIs were evaluated in one model. Indoor public gathering bans with a maximum of 10 or fewer were classified as severe. Indoor public gathering bans with maximums greater than 10 were classified as mild. Indoor restaurant dining ban was defined as adopted when indoor dining was banned and discontinued when indoor dining was reinstated, regardless of capacity specification or outdoor dining policies.

### NPI influence on COVID-19 Death Velocities

A total of 461 death breakpoints were identified (**Table 1**). 330 (71.6%) of breakpoints corresponded to a decrease in death velocity and 131 (28.4%) corresponded to an increase in death velocity. Across all 50 states, the median number of death breakpoints was 9 (minimum of 4 and maximum of 17) (**Figure S3**). Cumulative deaths over time were plotted along with their estimated breakpoints for the three best and worst performing states by cases per capita (**Figure S6**). Plots of the derivative of the logarithm of cumulative deaths (velocity) and the logarithm of the derivative of the logarithm (data implemented in modeling) for these states is available in the supplementary material (**Figures S7 and S8**).

Ordinal logistic regression models including a single policy estimated significantly increased odds of a decrease in death velocity following adoption of stay at home orders (OR 2.00, 95% CI 1.53-2.62), indoor restaurant dining ban (OR 1.50, 95% CI 1.15-1.95), and severe indoor public gathering bans (OR 1.45, 95% CI 1.12-1.88) (**Table 2**). In mutually adjusted models, institution of a stay at home order was associated with decreasing death velocity (AOR 1.89, 95% CI 1.25-2.87) (**Table 3**).

## DISCUSSION

We found multiple NPIs associated with decreasing COVID-19 case and mortality burdens in the U.S. With respect to cases, our adjusted time-dependent models found that stay at home orders and public mask mandates were effective at decreasing the rate of new diagnoses of COVID-19. Public mask mandates were associated with over twice the likelihood of reduced COVID-19 transmission even after adjusting for other policies that may have been adopted concurrently. Public mask mandates may encourage behavioral modifications as well as directly reduce the odds of transmission by using a physical barrier.^27,28^ COVID-19 is now understood to be transmitted primarily through aerosol spread in close contact.^29^ The state level observations provide support for masking mandates in reducing the case burden of respiratory epidemics or pandemics.

States that adopted mild indoor gathering bans had increases in the COVID-19 case burdens relative to states that did not adopt a mild indoor public gathering ban. Results from the mutually adjusted policy model suggested indoor restaurant dining bans and severe indoor public gathering may be associated with decreased case velocity (both *p* < .10), but more research is needed. Overall, it appears that gathering bans with limits greater than 10 were insufficient or exacerbated COVID-19 spread. This may be because these bans were often selected as an alternative to severe bans, which is suggested to be a more effective NPI by our findings. These observations are consistent with concerns regarding the indoor transmission of COVID-19 among large groups of individuals in public settings. NPI policies that discourage large gatherings are effective at reducing respiratory transmission.

With respect to mortality, the only NPI associated with decreasing COVID-19 mortality was stay at home order. Although other NPIs had a suggestion for benefit, our modeling approach was unable to detect significant associations with mortality. Given sample size limitations, limited variation in some policy adoption, and temporal variation in the progression of COVID-19 to death, we are limited in our ability to attribute deviations in daily death counts to specific policy actions. An additional consideration is that NPIs associated with decreasing case velocities but not associated with decreasing subsequent deaths may signal that case decreases occurred disproportionally among younger individuals with less risk for COVID-19 mortality. This may be particularly true for public mask mandates, which were significantly associated with decreased case but not mortality burden in adjusted models.

Our modeling approach allowed us to evaluate the merits of various NPIs concomitantly in a time-dependent fashion. Prior NPI studies have generally focused on pandemic influenza and relied on expert opinion or modeling rather than real-world data.^7,9,15,30,31^ In fact, the most recent Pandemic Influenza Plan by the U.S. Department of Health and Human Services described study of NPIs in the status of a data collection phase.^15^ Some retrospective data regarding NPIs and viral pandemics has been published. An analysis of U.S. cities found an association between increased duration of NPIs and total mortality reduction.^30^ Auger et al^13^ found school closures were associated with decreased COVID-19 incidence and mortality but adjustment for other NPIs was not included. Bendavid et al^32^ reported, in an international comparison of 10 countries including the U.S., no observable benefit of more restrictive NPIs (stay at home order, nonessential business closures) compared to less restrictive NPIs (social distancing guidelines, discouraging travel, and ban on large gatherings). We find the limited sample size and lack of variation in this study makes an absence of evidence conclusion difficult. Our analysis found both the strength and the direction of benefit related to severity of a gathering ban matters when assessing transmission dynamics. We found multiple NPIs to be associated with decreased case burden of COVID-19 in adjusted models (stay at home, public masking mandate, and severe gathering ban), which is supportive of prior expert opinions encouraging early, sustained, and layered application of NPIs to mitigate consequences of pandemic viral disease.

Our analyses include several limitations. Recommendations and policies were variably enforced by state governments. Populations in the U.S. may have had variable responses to stated policies based on preferences and beliefs. We did not attempt to measure markers of behavioral change based on the institution of policies and focused primarily on outcomes such as known transmission and death outcomes. We smoothed daily case counts by state and used a breakpoint analysis to cumulatively analyze distinct periods of times characterized by similar case velocities. This approach allowed for aggregating unique periods of COVID-19 burden across the limited sample of states (N=50). Our model does not account for national recommendations and policies for the entire U.S. population. The two most prominent of these announcements included the CDC recommendation of wearing cloth face coverings in public beginning April 3^rd^ 2020^4^ and “The President’s Coronavirus Guidelines for America” enacted March 16^th^ 2020 which included avoiding non-essential travel and avoiding social gatherings in groups of more than 10 people.^3^ Additionally, this study does not account for county or municipal level variation in NPI policies. Differences in testing capacity between states, especially early in the pandemic, were not accounted for in this analysis. Underreporting of cases may have biased our results towards the null. This analysis is unable to evaluate the impact of NPIs with substantial temporal overlap, such as K-12 school closures, and the impact of multiple NPIs adopted during concomitant periods may contribute to instability of estimates. To address this limitation, a smaller subset of NPIs were selected for evaluation.

Our model also relies on several unverified assumptions, such as the length and placement of the policy adoption window relative to a given week, and the 2-week minimal segment specification for breakpoint identification. Although these decisions were based on expert knowledge and review of the literature, the impact of these assumptions is unknown. Furthermore, any uncertainty in the establishment of the empirically estimated breakpoints was not reflected in the subsequent policy models which could suggest less precision of the final estimates we report.

In conclusion, adoption of several NPIs employed by U.S. states during the COVID-19 pandemic were associated with subsequent decreases in COVID-19 case burden. When accounting for the adoption of several NPIs, stay at home order was the NPI most strongly associated with decreases in COVID-19 mortality. Both restaurant dining and severe indoor public gathering bans (less than 10 people), were more effective in reducing transmission compared to mild indoor gathering bans (greater than 10). These findings reinforce efforts to deploy NPIs early and encourage adherence to limit the spread of dangerous respiratory epidemics.

## Data Availability

Data referred to in the manuscript can be made available by request at the discretion of the authors.

## ABBREVIATIONS

(AOR): Adjusted odds ratio
(COVID-19): Coronavirus disease 2019
(CSTE): Council of State and Territorial Epidemiologists
(GEE): Generalized estimating equations
(IFR): Infection fatality ratio
(NPI): Non-pharmaceutical intervention
(PCR): Polymerase chain reaction
(SARS): Severe acute respiratory syndrome
(SARS-CoV-2): Severe acute respiratory syndrome coronavirus 2

## Acknowledgements

None

