## Supplemental Material for "Non-Pharmaceutical Interventions and COVID-19 Burden in the United States"

**Supplementary Material**

### COVID-19 Natural History Estimations

#### *Estimated Time of COVID-19 Infection to Diagnosis*

We used a range 5 to 10 days to estimate time from COVID-19 infection to diagnosis of the case by community laboratory testing. In doing so, we estimated both the time from infection to symptom onset, as well as time from symptom onset to laboratory diagnosis. We estimated the time from COVID-19 infection to symptom onset to be a median of 5 days with interquartile range of 4 to 7 based on aggregate analysis of multiple peer-reviewed publications of confirmed COVID-19 cases in China.[1–7] We then estimated the time from COVID-19 symptom onset to laboratory diagnosis to be a median of 2 days with interquartile range of 1 to 3, factoring in data reported from China[1,3] and limitations in U.S. testing.[8] We also considered that the time from symptom onset to laboratory diagnosis would likely decrease over the duration of the pandemic, based on data describing COVID-19 in China[4,5] and data describing reductions in this time during the severe acute respiratory syndrome epidemic in Hong Kong.[3] Combining the estimations of COVID-19 infection to symptoms and symptoms to diagnosis yield an interquartile range of 5 to 10 days.

#### *Estimated Time of COVID-19 Diagnosis to Death*

We used an *a priori* definition of range 6 to 15 days to estimate time from COVID-19 diagnosis to death. This definition was informed by data reported by the Centers for Disease Control and Prevention describing patients who died with COVID-19 in the United States from February 12, 2020 to May 18, 2020.[9] Review of literature reporting time from illness onset to death in China was grossly similar although generally greater time periods.[3,8,10]

**Model Phasing**

Phases were aligned with calendar weeks being modeled. These phases were determined subjectively based on total numbers of COVID-19 cases over different periods of the pandemic. Four were used in the presented modeling: (1) March 22, 2020 – June 13, 2020, (2) June 14, 2020 – September 12, 2020, (3) September 13, 2020 – December 12, 2020, and (4) December 12, 2020 – February 12, 2021. These phases agree with chronological patterns in the type of breakpoint (increase vs. decrease in case velocity) over the study period (**Figures S2 and S3**) but are an unverifiable assumption in our models.

**Table S1. Dates of Non-Pharmaceutical Intervention Adoptions and Discontinuations by U.S. State**

| *State* | *Policy* | *Gathering Ban Level* | *Date* |
|---|---|---|---|
| **Alabama** | indoor restaurant dining ban (on) | | 03/17/2020 |
| | indoor public gathering ban | mild | 03/19/2020 |
| | indoor public gathering ban | severe | 03/28/2020 |
| | stay at home order (on) | | 04/04/2020 |
| | stay at home order (off) | | 04/30/2020 |
| | indoor public gathering ban | no gathering ban | 05/11/2020 |
| | indoor restaurant dining ban (off) | | 05/11/2020 |
| | public mask mandate (on) | | 07/16/2020 |
| **Alaska** | indoor restaurant dining ban (on) | | 03/18/2020 |
| | indoor public gathering ban | severe | 03/24/2020 |
| | stay at home order (on) | | 03/28/2020 |
| | stay at home order (off) | | 04/24/2020 |
| | indoor restaurant dining ban (off) | | 05/19/2020 |
| | indoor public gathering ban | no gathering ban | 05/22/2020 |
| **Arizona** | indoor public gathering ban | severe | 03/16/2020 |
| | indoor restaurant dining ban (on) | | 03/31/2020 |
| | stay at home order (on) | | 03/31/2020 |
| | indoor restaurant dining ban (off) | | 05/11/2020 |
| | indoor public gathering ban | no gathering ban | 05/12/2020 |
| | stay at home order (off) | | 05/15/2020 |
| | indoor public gathering ban | mild | 06/29/2020 |
| **Arkansas** | indoor restaurant dining ban (on) | | 03/19/2020 |
| | indoor public gathering ban | severe | 03/27/2020 |
| | indoor public gathering ban | mild | 05/04/2020 |
| | indoor restaurant dining ban (off) | | 05/11/2020 |
| | public mask mandate (on) | | 07/20/2020 |
| | indoor public gathering ban | severe | 01/02/2021 |
| **California** | indoor public gathering ban | mild | 03/11/2020 |
| | indoor public gathering ban | severe | 03/19/2020 |
| | indoor restaurant dining ban (on) | | 03/19/2020 |
| | stay at home order (on) | | 03/19/2020 |
| | indoor restaurant dining ban (off) | | 05/19/2020 |
| | public mask mandate (on) | | 06/18/2020 |
| | indoor restaurant dining ban (on) | | 07/13/2020 |

| | | | |
|---|---|---|---|
| | indoor restaurant dining ban (off) | | 08/28/2020 |
| | stay at home order (off) | | 01/25/2021 |
| **Colorado** | indoor public gathering ban | mild | 03/13/2020 |
| | indoor restaurant dining ban (on) | | 03/17/2020 |
| | indoor public gathering ban | severe | 03/19/2020 |
| | stay at home order (on) | | 03/26/2020 |
| | stay at home order (off) | | 04/26/2020 |
| | indoor restaurant dining ban (off) | | 05/27/2020 |
| | public mask mandate (on) | | 07/16/2020 |
| **Connecticut** | indoor public gathering ban | mild | 03/12/2020 |
| | indoor restaurant dining ban (on) | | 03/16/2020 |
| | stay at home order (on) | | 03/23/2020 |
| | indoor public gathering ban | severe | 03/26/2020 |
| | public mask mandate (on) | | 04/20/2020 |
| | stay at home order (off) | | 05/20/2020 |
| | indoor public gathering ban | mild | 06/17/2020 |
| | indoor restaurant dining ban (off) | | 06/17/2020 |
| **Delaware** | indoor public gathering ban | mild | 03/16/2020 |
| | indoor restaurant dining ban (on) | | 03/16/2020 |
| | stay at home order (on) | | 03/24/2020 |
| | indoor public gathering ban | severe | 04/02/2020 |
| | public mask mandate (on) | | 04/28/2020 |
| | stay at home order (off) | | 05/31/2020 |
| | indoor restaurant dining ban (off) | | 06/01/2020 |
| | indoor public gathering ban | mild | 06/15/2020 |
| | indoor public gathering ban | severe | 12/14/2020 |
| **Florida** | indoor public gathering ban | mild | 03/12/2020 |
| | indoor restaurant dining ban (on) | | 03/20/2020 |
| | indoor public gathering ban | severe | 04/03/2020 |
| | stay at home order (on) | | 04/03/2020 |
| | stay at home order (off) | | 05/04/2020 |
| | indoor restaurant dining ban (off) | | 05/18/2020 |
| | indoor public gathering ban | mild | 06/05/2020 |
| | indoor public gathering ban | no gathering ban | 09/25/2020 |
| **Georgia** | indoor public gathering ban | mild | 03/19/2020 |
| | indoor public gathering ban | severe | 03/23/2020 |
| | indoor restaurant dining ban (on) | | 04/03/2020 |

| | | | |
|---|---|---|---|
| | stay at home order (on) | | 04/03/2020 |
| | indoor restaurant dining ban (off) | | 04/27/2020 |
| | stay at home order (off) | | 04/30/2020 |
| | indoor public gathering ban | mild | 06/01/2020 |
| **Hawaii** | indoor restaurant dining ban (on) | | 03/17/2020 |
| | indoor public gathering ban | severe | 03/25/2020 |
| | stay at home order (on) | | 03/25/2020 |
| | public mask mandate (on) | | 04/25/2020 |
| | stay at home order (off) | | 05/31/2020 |
| | indoor restaurant dining ban (off) | | 06/05/2020 |
| | indoor restaurant dining ban (on) | | 08/27/2020 |
| | stay at home order (on) | | 08/27/2020 |
| | indoor restaurant dining ban (off) | | 09/24/2020 |
| | stay at home order (off) | | 09/24/2020 |
| **Idaho** | indoor public gathering ban | mild | 03/16/2020 |
| | indoor public gathering ban | severe | 03/19/2020 |
| | indoor restaurant dining ban (on) | | 03/25/2020 |
| | stay at home order (on) | | 03/25/2020 |
| | stay at home order (off) | | 04/30/2020 |
| | indoor restaurant dining ban (off) | | 05/16/2020 |
| **Illinois** | indoor public gathering ban | mild | 03/12/2020 |
| | indoor restaurant dining ban (on) | | 03/16/2020 |
| | indoor public gathering ban | severe | 03/21/2020 |
| | stay at home order (on) | | 03/21/2020 |
| | public mask mandate (on) | | 05/01/2020 |
| | stay at home order (off) | | 05/29/2020 |
| | indoor public gathering ban | mild | 06/26/2020 |
| | indoor restaurant dining ban (off) | | 06/26/2020 |
| | indoor restaurant dining ban (on) | | 10/22/2020 |
| | indoor public gathering ban | severe | 11/20/2020 |
| | indoor public gathering ban | mild | 01/15/2021 |
| | indoor restaurant dining ban (off) | | 01/19/2021 |
| **Indiana** | indoor public gathering ban | mild | 03/12/2020 |
| | indoor public gathering ban | severe | 03/16/2020 |
| | indoor restaurant dining ban (on) | | 03/16/2020 |
| | stay at home order (on) | | 03/24/2020 |
| | stay at home order (off) | | 05/01/2020 |

| | | | |
|---|---|---|---|
| | indoor public gathering ban | mild | 05/04/2020 |
| | indoor restaurant dining ban (off) | | 05/11/2020 |
| | public mask mandate (on) | | 07/27/2020 |
| | indoor public gathering ban | no gathering ban | 09/26/2020 |
| **Iowa** | indoor public gathering ban | severe | 03/17/2020 |
| | indoor restaurant dining ban (on) | | 03/17/2020 |
| | indoor restaurant dining ban (off) | | 05/15/2020 |
| | indoor public gathering ban | mild | 06/01/2020 |
| | public mask mandate (on) | | 11/17/2020 |
| **Kansas** | indoor public gathering ban | mild | 03/16/2020 |
| | indoor public gathering ban | severe | 03/25/2020 |
| | indoor restaurant dining ban (on) | | 03/30/2020 |
| | stay at home order (on) | | 03/30/2020 |
| | stay at home order (off) | | 05/03/2020 |
| | indoor restaurant dining ban (off) | | 05/04/2020 |
| | indoor public gathering ban | mild | 05/22/2020 |
| | public mask mandate (on) | | 07/03/2020 |
| **Kentucky** | indoor restaurant dining ban (on) | | 03/16/2020 |
| | indoor public gathering ban | mild | 03/19/2020 |
| | indoor public gathering ban | severe | 03/25/2020 |
| | stay at home order (on) | | 03/26/2020 |
| | indoor restaurant dining ban (off) | | 05/22/2020 |
| | indoor public gathering ban | mild | 06/29/2020 |
| | stay at home order (off) | | 06/29/2020 |
| | public mask mandate (on) | | 07/09/2020 |
| | indoor public gathering ban | severe | 07/20/2020 |
| **Louisiana** | indoor public gathering ban | mild | 03/12/2020 |
| | indoor restaurant dining ban (on) | | 03/16/2020 |
| | indoor public gathering ban | severe | 03/22/2020 |
| | stay at home order (on) | | 03/23/2020 |
| | indoor public gathering ban | mild | 05/15/2020 |
| | indoor restaurant dining ban (off) | | 05/15/2020 |
| | stay at home order (off) | | 05/15/2020 |
| | public mask mandate (on) | | 07/13/2020 |
| **Maine** | indoor public gathering ban | mild | 03/12/2020 |
| | indoor public gathering ban | severe | 03/18/2020 |
| | indoor restaurant dining ban (on) | | 03/18/2020 |

| | | | |
|---|---|---|---|
| | stay at home order (on) | | 04/02/2020 |
| | public mask mandate (on) | | 05/01/2020 |
| | indoor restaurant dining ban (off) | | 05/18/2020 |
| | indoor public gathering ban | mild | 05/31/2020 |
| | stay at home order (off) | | 05/31/2020 |
| **Maryland** | indoor public gathering ban | mild | 03/12/2020 |
| | indoor restaurant dining ban (on) | | 03/16/2020 |
| | indoor public gathering ban | severe | 03/19/2020 |
| | stay at home order (on) | | 03/30/2020 |
| | public mask mandate (on) | | 04/18/2020 |
| | stay at home order (off) | | 05/15/2020 |
| | indoor restaurant dining ban (off) | | 06/12/2020 |
| | indoor public gathering ban | no gathering ban | 09/01/2020 |
| | indoor public gathering ban | severe | 12/17/2020 |
| **Massachusetts** | indoor public gathering ban | mild | 03/13/2020 |
| | indoor restaurant dining ban (on) | | 03/17/2020 |
| | indoor public gathering ban | severe | 03/24/2020 |
| | stay at home order (on) | | 03/24/2020 |
| | public mask mandate (on) | | 05/06/2020 |
| | stay at home order (off) | | 05/18/2020 |
| | indoor restaurant dining ban (off) | | 06/22/2020 |
| | indoor public gathering ban | mild | 07/06/2020 |
| | indoor public gathering ban | severe | 11/06/2020 |
| **Michigan** | indoor public gathering ban | mild | 03/13/2020 |
| | indoor restaurant dining ban (on) | | 03/16/2020 |
| | indoor public gathering ban | severe | 03/24/2020 |
| | stay at home order (on) | | 03/24/2020 |
| | indoor restaurant dining ban (off) | | 05/22/2020 |
| | stay at home order (off) | | 06/01/2020 |
| | indoor public gathering ban | mild | 06/10/2020 |
| | public mask mandate (on) | | 07/10/2020 |
| | public mask mandate (off) | | 10/02/2020 |
| | public mask mandate (on) | | 10/05/2020 |
| | indoor restaurant dining ban (on) | | 12/09/2020 |
| | indoor public gathering ban | severe | 12/21/2020 |
| **Minnesota** | indoor public gathering ban | mild | 03/13/2020 |
| | indoor restaurant dining ban (on) | | 03/17/2020 |

| | | | |
|---|---|---|---|
| | indoor public gathering ban | severe | 03/26/2020 |
| | stay at home order (on) | | 03/27/2020 |
| | stay at home order (off) | | 05/17/2020 |
| | indoor public gathering ban | mild | 05/18/2020 |
| | indoor restaurant dining ban (off) | | 06/05/2020 |
| | public mask mandate (on) | | 07/25/2020 |
| | indoor restaurant dining ban (on) | | 11/21/2020 |
| | indoor restaurant dining ban (off) | | 01/11/2021 |
| | indoor public gathering ban | severe | 01/13/2021 |
| **Mississippi** | indoor public gathering ban | severe | 03/25/2020 |
| | indoor restaurant dining ban (on) | | 04/03/2020 |
| | stay at home order (on) | | 04/03/2020 |
| | stay at home order (off) | | 04/27/2020 |
| | indoor restaurant dining ban (off) | | 05/07/2020 |
| | indoor public gathering ban | mild | 06/01/2020 |
| | indoor public gathering ban | severe | 07/13/2020 |
| | public mask mandate (on) | | 08/05/2020 |
| | public mask mandate (off) | | 09/30/2020 |
| | public mask mandate (on) | | 10/19/2020 |
| **Missouri** | indoor public gathering ban | mild | 03/15/2020 |
| | indoor public gathering ban | severe | 03/23/2020 |
| | indoor restaurant dining ban (on) | | 04/06/2020 |
| | stay at home order (on) | | 04/06/2020 |
| | stay at home order (off) | | 05/03/2020 |
| | indoor public gathering ban | no gathering ban | 05/04/2020 |
| | indoor restaurant dining ban (off) | | 05/04/2020 |
| **Montana** | indoor public gathering ban | mild | 03/16/2020 |
| | indoor restaurant dining ban (on) | | 03/20/2020 |
| | indoor public gathering ban | severe | 03/24/2020 |
| | stay at home order (on) | | 03/28/2020 |
| | stay at home order (off) | | 04/26/2020 |
| | indoor restaurant dining ban (off) | | 05/04/2020 |
| | indoor public gathering ban | mild | 06/01/2020 |
| | public mask mandate (on) | | 11/17/2020 |
| | indoor public gathering ban | no gathering ban | 01/15/2021 |
| **Nebraska** | indoor public gathering ban | mild | 03/13/2020 |
| | indoor public gathering ban | severe | 03/16/2020 |

| | | | |
|---|---|---|---|
| | indoor restaurant dining ban (on) | | 03/19/2020 |
| | indoor restaurant dining ban (off) | | 05/04/2020 |
| | indoor public gathering ban | mild | 06/01/2020 |
| **Nevada** | indoor restaurant dining ban (on) | | 03/20/2020 |
| | indoor public gathering ban | severe | 03/24/2020 |
| | stay at home order (on) | | 03/31/2020 |
| | indoor restaurant dining ban (off) | | 05/09/2020 |
| | stay at home order (off) | | 05/15/2020 |
| | indoor public gathering ban | mild | 05/29/2020 |
| | public mask mandate (on) | | 06/26/2020 |
| **New Hampshire** | indoor public gathering ban | mild | 03/16/2020 |
| | indoor restaurant dining ban (on) | | 03/16/2020 |
| | indoor public gathering ban | severe | 03/24/2020 |
| | stay at home order (on) | | 03/27/2020 |
| | indoor public gathering ban | no gathering ban | 06/15/2020 |
| | indoor restaurant dining ban (off) | | 06/15/2020 |
| | stay at home order (off) | | 06/15/2020 |
| | public mask mandate (on) | | 11/20/2020 |
| **New Jersey** | indoor public gathering ban | mild | 03/16/2020 |
| | indoor public gathering ban | severe | 03/21/2020 |
| | indoor restaurant dining ban (on) | | 03/21/2020 |
| | stay at home order (on) | | 03/21/2020 |
| | stay at home order (off) | | 06/09/2020 |
| | indoor public gathering ban | mild | 06/22/2020 |
| | public mask mandate (on) | | 07/08/2020 |
| | indoor restaurant dining ban (off) | | 09/04/2020 |
| | indoor public gathering ban | severe | 12/07/2020 |
| **New Mexico** | indoor public gathering ban | mild | 03/12/2020 |
| | indoor restaurant dining ban (on) | | 03/19/2020 |
| | indoor public gathering ban | severe | 03/24/2020 |
| | stay at home order (on) | | 03/24/2020 |
| | public mask mandate (on) | | 05/16/2020 |
| | indoor restaurant dining ban (off) | | 06/01/2020 |
| | stay at home order (off) | | 06/30/2020 |
| | indoor restaurant dining ban (on) | | 07/13/2020 |
| | indoor restaurant dining ban (off) | | 08/29/2020 |
| | stay at home order (on) | | 11/13/2020 |

| | | | |
|---|---|---|---|
| | indoor restaurant dining ban (on) | | 11/16/2020 |
| | stay at home order (off) | | 11/30/2020 |
| | indoor restaurant dining ban (off) | | 12/02/2020 |
| **New York** | indoor public gathering ban | mild | 03/12/2020 |
| | indoor restaurant dining ban (on) | | 03/16/2020 |
| | indoor public gathering ban | severe | 03/22/2020 |
| | stay at home order (on) | | 03/22/2020 |
| | public mask mandate (on) | | 04/17/2020 |
| | stay at home order (off) | | 05/28/2020 |
| | indoor restaurant dining ban (off) | | 06/12/2020 |
| | indoor public gathering ban | mild | 06/15/2020 |
| | indoor public gathering ban | severe | 11/11/2020 |
| **North Carolina** | indoor public gathering ban | mild | 03/14/2020 |
| | indoor restaurant dining ban (on) | | 03/17/2020 |
| | indoor public gathering ban | severe | 03/30/2020 |
| | stay at home order (on) | | 03/30/2020 |
| | indoor restaurant dining ban (off) | | 05/22/2020 |
| | stay at home order (off) | | 05/22/2020 |
| | public mask mandate (on) | | 06/26/2020 |
| **North Dakota** | indoor restaurant dining ban (on) | | 03/20/2020 |
| | indoor restaurant dining ban (off) | | 05/01/2020 |
| | public mask mandate (on) | | 11/14/2020 |
| | public mask mandate (off) | | 01/18/2021 |
| **Ohio** | indoor public gathering ban | mild | 03/12/2020 |
| | indoor restaurant dining ban (on) | | 03/15/2020 |
| | indoor public gathering ban | severe | 03/22/2020 |
| | stay at home order (on) | | 03/23/2020 |
| | stay at home order (off) | | 05/19/2020 |
| | indoor restaurant dining ban (off) | | 05/21/2020 |
| | public mask mandate (on) | | 07/23/2020 |
| **Oklahoma** | indoor public gathering ban | severe | 03/24/2020 |
| | indoor restaurant dining ban (on) | | 03/25/2020 |
| | indoor restaurant dining ban (off) | | 05/01/2020 |
| | indoor public gathering ban | mild | 05/15/2020 |
| | indoor public gathering ban | mild | 12/10/2020 |
| **Oregon** | indoor public gathering ban | mild | 03/12/2020 |
| | indoor restaurant dining ban (on) | | 03/17/2020 |

| | | | |
|---|---|---|---|
| | indoor public gathering ban | severe | 03/23/2020 |
| | stay at home order (on) | | 03/23/2020 |
| | indoor public gathering ban | mild | 05/15/2020 |
| | indoor restaurant dining ban (off) | | 05/15/2020 |
| | stay at home order (off) | | 06/19/2020 |
| | public mask mandate (on) | | 07/01/2020 |
| | indoor public gathering ban | severe | 07/15/2020 |
| | indoor restaurant dining ban (on) | | 11/18/2020 |
| | indoor restaurant dining ban (off) | | 12/02/2020 |
| | indoor public gathering ban | mild | 12/18/2020 |
| **Pennsylvania** | indoor restaurant dining ban (on) | | 03/16/2020 |
| | indoor public gathering ban | severe | 03/17/2020 |
| | stay at home order (on) | | 04/01/2020 |
| | indoor public gathering ban | mild | 05/08/2020 |
| | indoor restaurant dining ban (off) | | 05/29/2020 |
| | stay at home order (off) | | 06/04/2020 |
| | public mask mandate (on) | | 07/01/2020 |
| | indoor public gathering ban | no gathering ban | 09/22/2020 |
| | indoor public gathering ban | mild | 10/01/2020 |
| | indoor restaurant dining ban (on) | | 12/12/2020 |
| | indoor restaurant dining ban (off) | | 01/04/2021 |
| **Rhode Island** | indoor public gathering ban | mild | 03/11/2020 |
| | indoor restaurant dining ban (on) | | 03/16/2020 |
| | indoor public gathering ban | severe | 03/28/2020 |
| | stay at home order (on) | | 03/28/2020 |
| | public mask mandate (on) | | 05/08/2020 |
| | stay at home order (off) | | 05/08/2020 |
| | indoor public gathering ban | mild | 06/01/2020 |
| | indoor restaurant dining ban (off) | | 06/01/2020 |
| **South Carolina** | indoor public gathering ban | mild | 03/15/2020 |
| | indoor restaurant dining ban (on) | | 03/17/2020 |
| | indoor public gathering ban | severe | 03/23/2020 |
| | stay at home order (on) | | 04/07/2020 |
| | stay at home order (off) | | 05/04/2020 |
| | indoor restaurant dining ban (off) | | 05/11/2020 |
| | indoor public gathering ban | mild | 06/11/2020 |
| **South Dakota** | indoor public gathering ban | severe | 03/23/2020 |

| | | | |
|---|---|---|---|
| | indoor public gathering ban | no gathering ban | 04/28/2020 |
| **Tennessee** | indoor public gathering ban | mild | 03/13/2020 |
| | indoor public gathering ban | severe | 03/22/2020 |
| | indoor restaurant dining ban (on) | | 03/23/2020 |
| | stay at home order (on) | | 03/31/2020 |
| | indoor restaurant dining ban (off) | | 04/27/2020 |
| | stay at home order (off) | | 04/30/2020 |
| | indoor public gathering ban | mild | 05/22/2020 |
| | indoor public gathering ban | no gathering ban | 09/29/2020 |
| | indoor public gathering ban | severe | 12/20/2020 |
| **Texas** | indoor restaurant dining ban (on) | | 03/20/2020 |
| | indoor public gathering ban | severe | 03/21/2020 |
| | stay at home order (on) | | 04/02/2020 |
| | stay at home order (off) | | 04/30/2020 |
| | indoor restaurant dining ban (off) | | 05/01/2020 |
| | indoor public gathering ban | mild | 05/05/2020 |
| | public mask mandate (on) | | 07/03/2020 |
| **Utah** | indoor public gathering ban | mild | 03/16/2020 |
| | indoor restaurant dining ban (on) | | 03/18/2020 |
| | indoor public gathering ban | severe | 03/21/2020 |
| | indoor restaurant dining ban (off) | | 05/01/2020 |
| | indoor public gathering ban | mild | 05/16/2020 |
| | public mask mandate (on) | | 11/08/2020 |
| | indoor public gathering ban | no gathering ban | 11/24/2020 |
| **Vermont** | indoor public gathering ban | mild | 03/13/2020 |
| | indoor restaurant dining ban (on) | | 03/17/2020 |
| | indoor public gathering ban | severe | 03/21/2020 |
| | stay at home order (on) | | 03/25/2020 |
| | stay at home order (off) | | 05/15/2020 |
| | indoor public gathering ban | mild | 06/01/2020 |
| | indoor restaurant dining ban (off) | | 06/05/2020 |
| | public mask mandate (on) | | 08/01/2020 |
| | indoor public gathering ban | severe | 11/14/2020 |
| **Virginia** | indoor public gathering ban | mild | 03/15/2020 |
| | indoor restaurant dining ban (on) | | 03/24/2020 |
| | indoor public gathering ban | severe | 03/25/2020 |
| | stay at home order (on) | | 03/30/2020 |

| | | | |
|---|---|---|---|
| | stay at home order (off) | | 05/28/2020 |
| | public mask mandate (on) | | 05/29/2020 |
| | indoor public gathering ban | mild | 06/05/2020 |
| | indoor restaurant dining ban (off) | | 06/05/2020 |
| | indoor public gathering ban | severe | 12/14/2020 |
| **Washington** | indoor public gathering ban | mild | 03/13/2020 |
| | indoor restaurant dining ban (on) | | 03/16/2020 |
| | indoor public gathering ban | severe | 03/23/2020 |
| | stay at home order (on) | | 03/23/2020 |
| | indoor restaurant dining ban (off) | | 05/11/2020 |
| | stay at home order (off) | | 05/31/2020 |
| | indoor public gathering ban | mild | 06/19/2020 |
| | public mask mandate (on) | | 06/26/2020 |
| | indoor public gathering ban | severe | 07/20/2020 |
| | indoor public gathering ban | mild | 10/13/2020 |
| | indoor restaurant dining ban (on) | | 11/18/2020 |
| | indoor restaurant dining ban (off) | | 01/28/2021 |
| **West Virginia** | indoor restaurant dining ban (on) | | 03/17/2020 |
| | indoor public gathering ban | severe | 03/24/2020 |
| | stay at home order (on) | | 03/24/2020 |
| | stay at home order (off) | | 05/03/2020 |
| | indoor restaurant dining ban (off) | | 05/21/2020 |
| | indoor public gathering ban | mild | 05/24/2020 |
| | public mask mandate (on) | | 07/07/2020 |
| **Wisconsin** | indoor public gathering ban | mild | 03/16/2020 |
| | indoor public gathering ban | severe | 03/17/2020 |
| | indoor restaurant dining ban (on) | | 03/17/2020 |
| | stay at home order (on) | | 03/25/2020 |
| | indoor public gathering ban | no gathering ban | 05/13/2020 |
| | indoor restaurant dining ban (off) | | 05/13/2020 |
| | stay at home order (off) | | 05/13/2020 |
| | public mask mandate (on) | | 08/01/2020 |
| | indoor public gathering ban | severe | 10/08/2020 |
| | indoor public gathering ban | no gathering ban | 11/10/2020 |
| **Wyoming** | indoor public gathering ban | mild | 03/13/2020 |
| | indoor restaurant dining ban (on) | | 03/19/2020 |
| | indoor public gathering ban | severe | 03/20/2020 |

| | | |
|---|---|---|
| indoor public gathering ban | mild | 05/15/2020 |
| indoor restaurant dining ban (off) | | 05/15/2020 |
| public mask mandate (on) | | 12/09/2020 |
| indoor public gathering ban | severe | 01/02/2021 |

### Table S2. Total Cases & Deaths & Per Capita (100,000) by U.S. State

| *State* | *Total Cases* | *Total Deaths* | *Cases Per Capita (100,000)* | *Deaths Per Capita (100,000)* |
|---|---|---|---|---|
| *Alabama* | 499,819 | 10,148 | 10,226 | 208 |
| *Alaska* | 56,886 | 305 | 7,714 | 41 |
| *Arizona* | 826,454 | 16,328 | 11,524 | 228 |
| *Arkansas* | 324,818 | 5,319 | 10,778 | 176 |
| *California* | 3,501,394 | 54,124 | 8,852 | 137 |
| *Colorado* | 436,602 | 5,989 | 7,666 | 105 |
| *Connecticut* | 285,330 | 7,704 | 7,986 | 216 |
| *Delaware* | 88,354 | 1,473 | 9,135 | 152 |
| *Florida* | 1,909,209 | 32,266 | 8,964 | 151 |
| *Georgia* | 1,023,487 | 17,906 | 9,729 | 170 |
| *Hawaii* | 28,699 | 445 | 2,020 | 31 |
| *Idaho* | 172,931 | 1,879 | 9,858 | 107 |
| *Illinois* | 1,198,335 | 23,014 | 9,405 | 181 |
| *Indiana* | 667,262 | 12,737 | 9,971 | 190 |
| *Iowa* | 282,384 | 5,558 | 8,947 | 176 |
| *Kansas* | 295,861 | 4,812 | 10,162 | 165 |
| *Kentucky* | 410,709 | 4,819 | 9,191 | 108 |
| *Louisiana* | 433,785 | 9,748 | 9,309 | 209 |
| *Maine* | 45,794 | 706 | 3,422 | 53 |
| *Maryland* | 387,319 | 7,955 | 6,410 | 132 |
| *Massachusetts* | 591,356 | 16,417 | 8,568 | 238 |
| *Michigan* | 656,072 | 16,658 | 6,563 | 167 |
| *Minnesota* | 490,011 | 6,550 | 8,733 | 117 |
| *Mississippi* | 297,581 | 6,808 | 9,964 | 228 |
| *Missouri* | 480,643 | 8,161 | 7,845 | 133 |
| *Montana* | 100,914 | 1,381 | 9,500 | 130 |
| *Nebraska* | 203,026 | 2,113 | 10,523 | 110 |
| *Nevada* | 296,190 | 5,037 | 9,761 | 166 |
| *New Hampshire* | 76,861 | 1,184 | 5,666 | 87 |
| *New Jersey* | 812,609 | 23,574 | 9,122 | 265 |
| *New Mexico* | 186,922 | 3,808 | 8,920 | 182 |
| *New York* | 1,681,169 | 39,029 | 8,603 | 200 |
| *North Carolina* | 872,176 | 11,502 | 8,400 | 111 |
| *North Dakota* | 100,391 | 1,478 | 13,208 | 194 |
| *Ohio* | 978,471 | 17,656 | 8,371 | 151 |
| *Oklahoma* | 428,997 | 4,534 | 10,880 | 115 |
| *Oregon* | 157,079 | 2,296 | 3,748 | 55 |
| *Pennsylvania* | 948,643 | 24,349 | 7,407 | 190 |
| *Rhode Island* | 128,781 | 2,547 | 12,180 | 241 |
| *South Carolina* | 525,865 | 8,754 | 10,343 | 172 |
| *South Dakota* | 113,589 | 1,900 | 12,875 | 215 |
| *Tennessee* | 782,206 | 11,543 | 11,554 | 171 |
| *Texas* | 2,686,818 | 44,451 | 9,361 | 155 |
| *Utah* | 378,850 | 1,976 | 11,985 | 63 |
| *Vermont* | 16,083 | 208 | 2,568 | 33 |
| *Virginia* | 585,700 | 9,596 | 6,876 | 113 |
| *Washington* | 344,532 | 5,041 | 4,572 | 67 |
| *West Virginia* | 133,445 | 2,325 | 7,390 | 129 |
| *Wisconsin* | 621,654 | 7,106 | 10,693 | 122 |
| *Wyoming* | 54,764 | 682 | 9,479 | 118 |

**Figure S1. Example Graphs of Alabama COVID-19 Case Data Manipulations for the Identification of Breakpoints.**

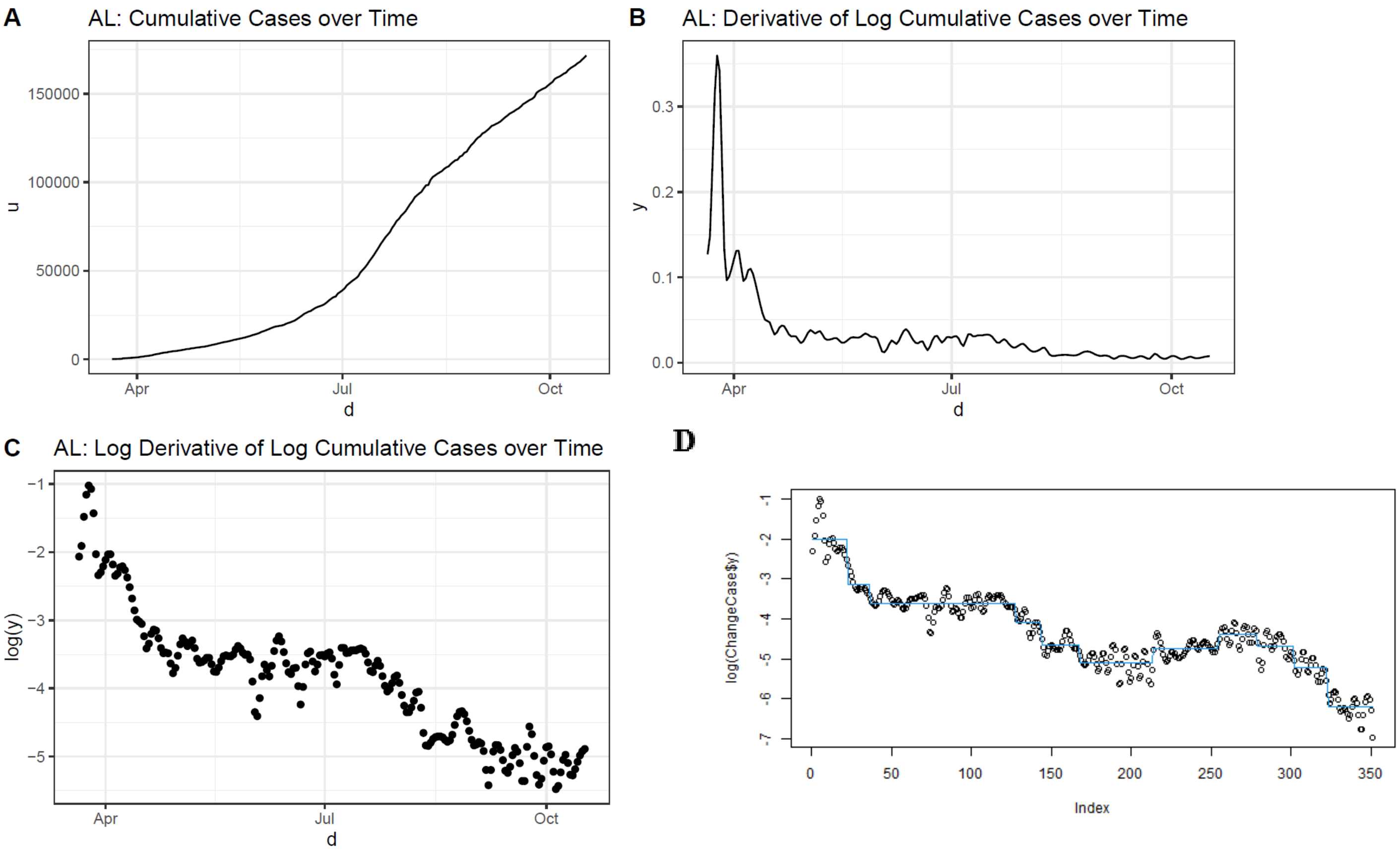

Figure S1(a) shows cumulative cases over time. Figure S1(b) shows the 1st derivative of the log of cumulative cases over time, or case velocity. Figure S1(c) shows the log of case velocity, which was mapped to an entire real line for modeling as a linear function in Figure S1(d). Based on this breakpoint approach, Alabama experienced eight decreases and two increases in case velocity.

**Figure S2. Count of State-Level Case Velocity Breakpoints by Week in the United States**

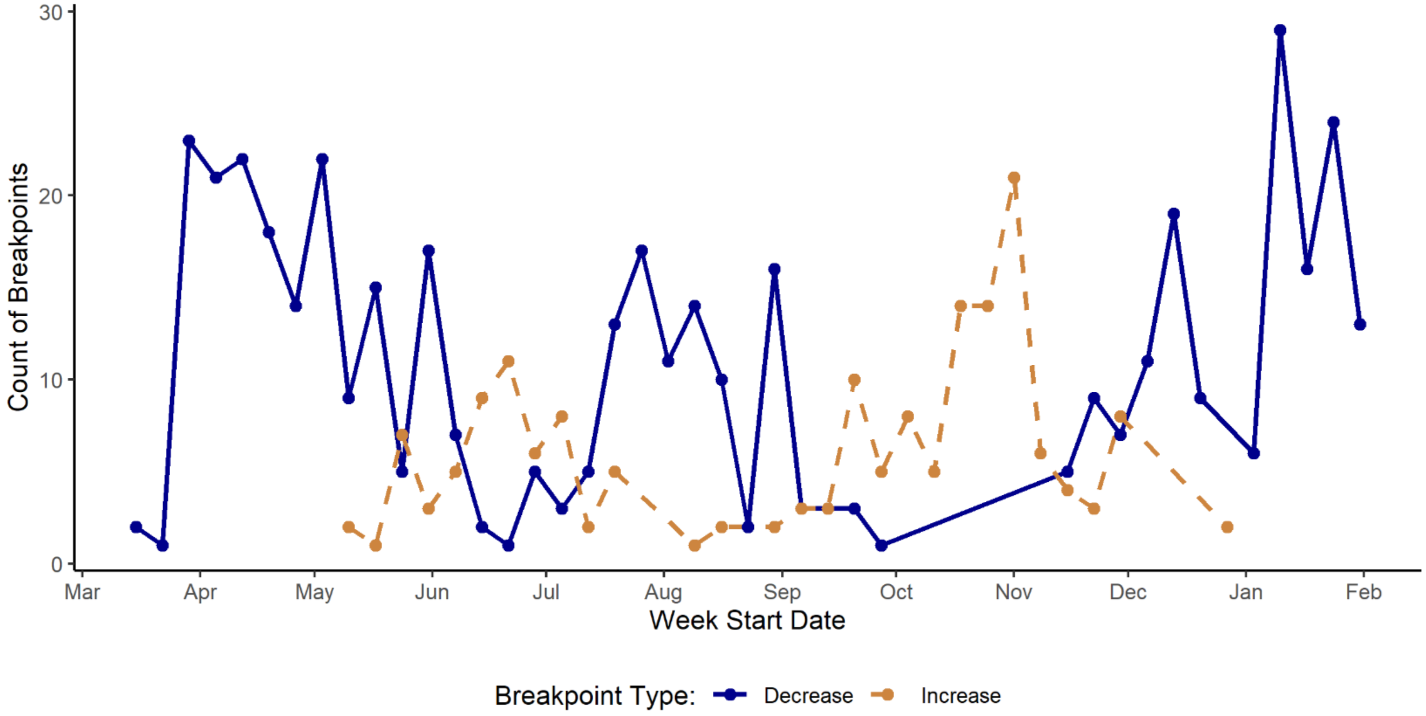

**Figure S3. Count of State-Level Death Velocity Breakpoints by Week in the United States**

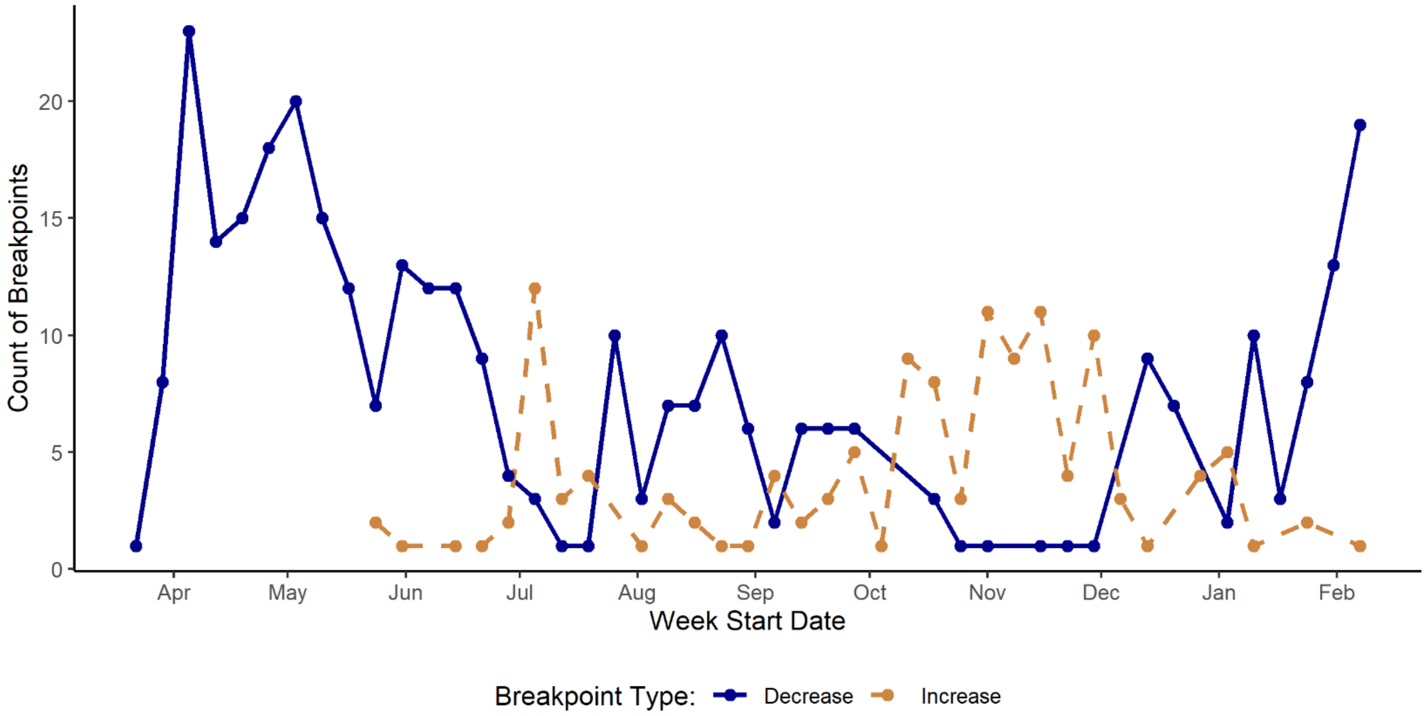

**Figure S4. Derivative of the logarithm of cumulative cases and the breakpoints identified for the top 3 lowest and highest U.S. states by COVID-19 cases per capita**

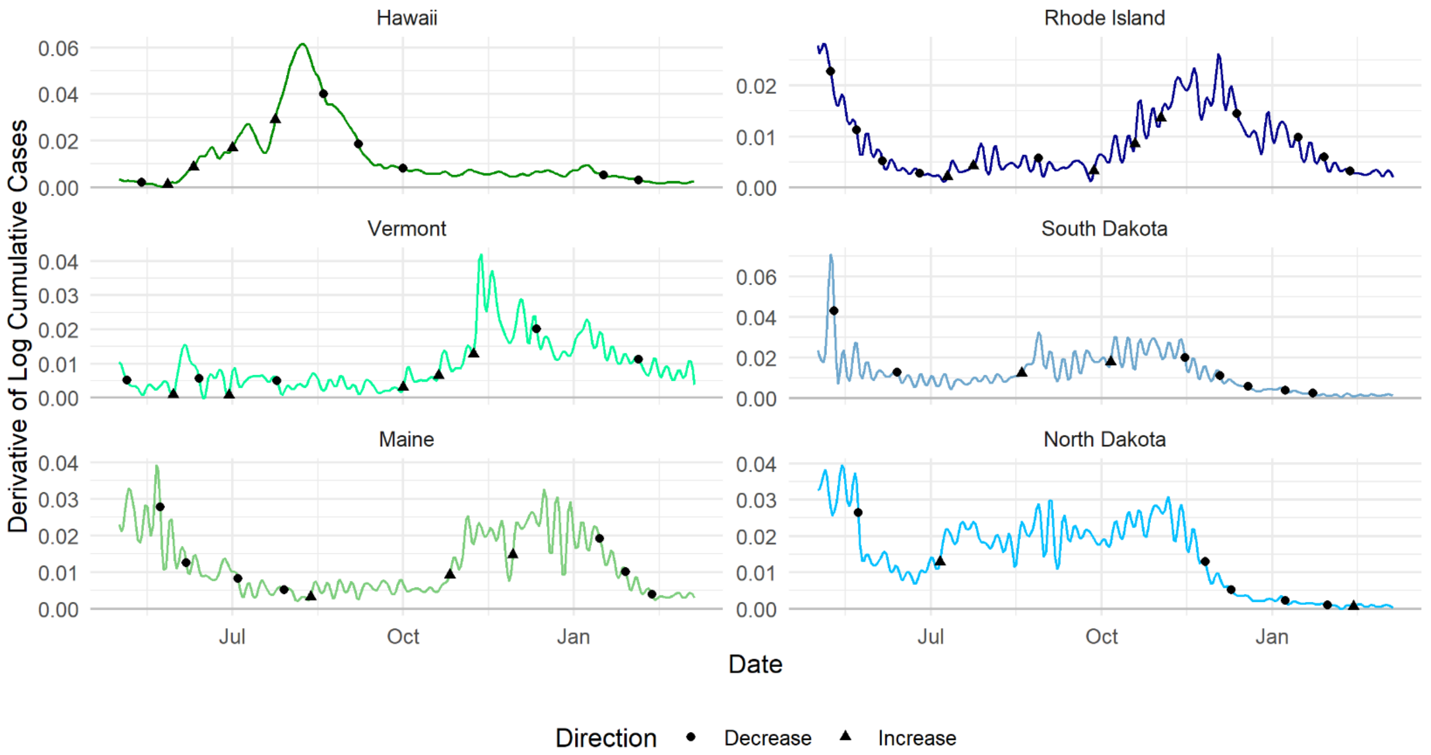

Derivative of the logarithm of cumulative cases most closely represents case velocities graphed. Y=0 for graphs were set as May 1st, 2020 rather than date of first cumulative case due to high initial velocities, pictorially obscuring clinically informative later breakpoints. Hawaii, Vermont, and Maine ranked as the three best performing states by cumulative number of COVID-19 cases per capita as of March 7, 2021. Rhode Island, South Dakota, and North Dakota ranked as the three worst performing states over the same time period. Breakpoints, dates at which the linear segments of COVID-19 case velocities showed substantial change in their rate, are plotted over the liner plot of cases for each respective state.

**Figure S5. Logarithm of the derivative of the logarithm of cumulative cases and the breakpoints identified for the top 3 lowest and highest U.S. states by COVID-19 cases per capita**

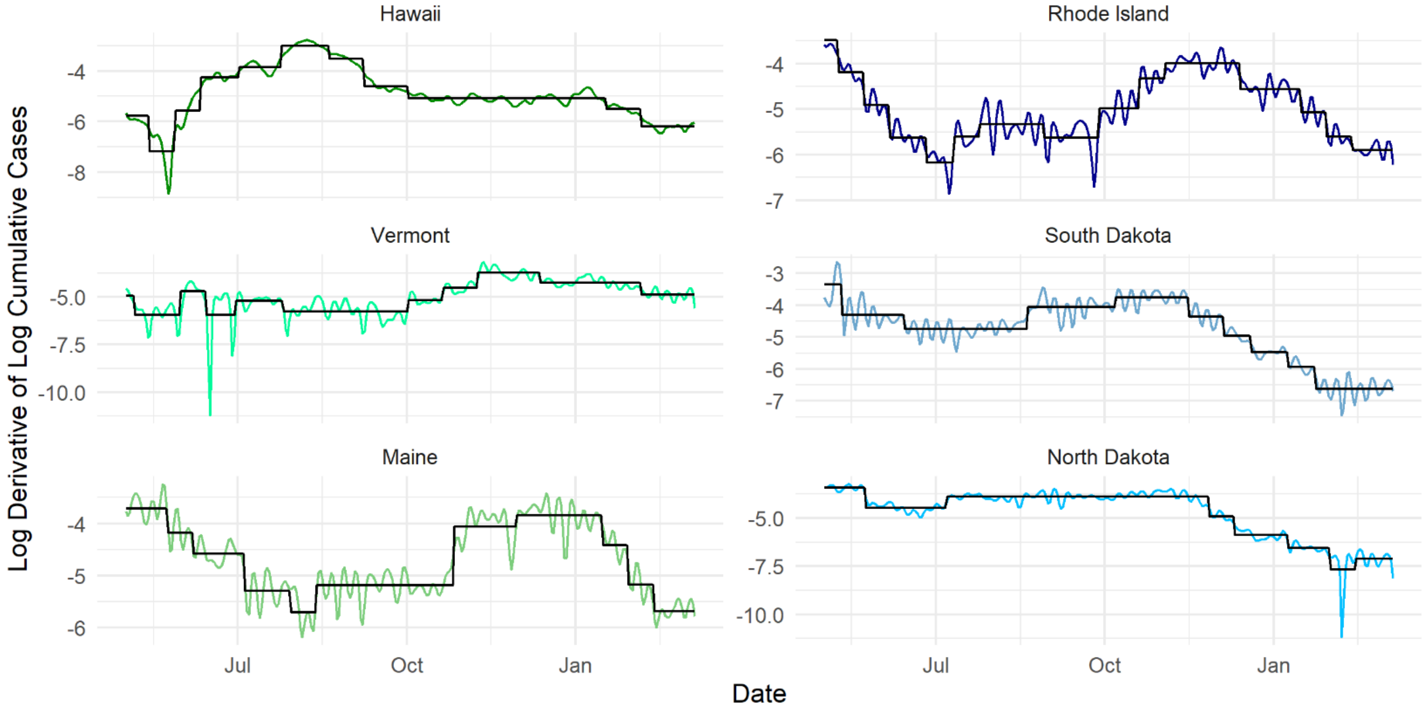

Logarithm of the derivative of the logarithm of cumulative cases most closely represents curation of data implemented to models. Y=0 for graphs were set as May 1st, 2020 rather than date of first cumulative case due to high initial velocities, pictorially obscuring clinically informative later breakpoints. Hawaii, Vermont, and Maine ranked as the three best performing states by cumulative number of COVID-19 cases per capita as of March 7, 2021. Rhode Island, South Dakota, and North Dakota ranked as the three worst performing states over the same time period. Breakpoints, dates at which the linear segments of COVID-19 case velocities showed substantial change in their rate, is fitted over the liner plot of cases for each respective state.

**Figure S6. Cumulative COVID-19 deaths and the breakpoints identified for the top 3 lowest and highest U.S. states by COVID-19 cases per capita**

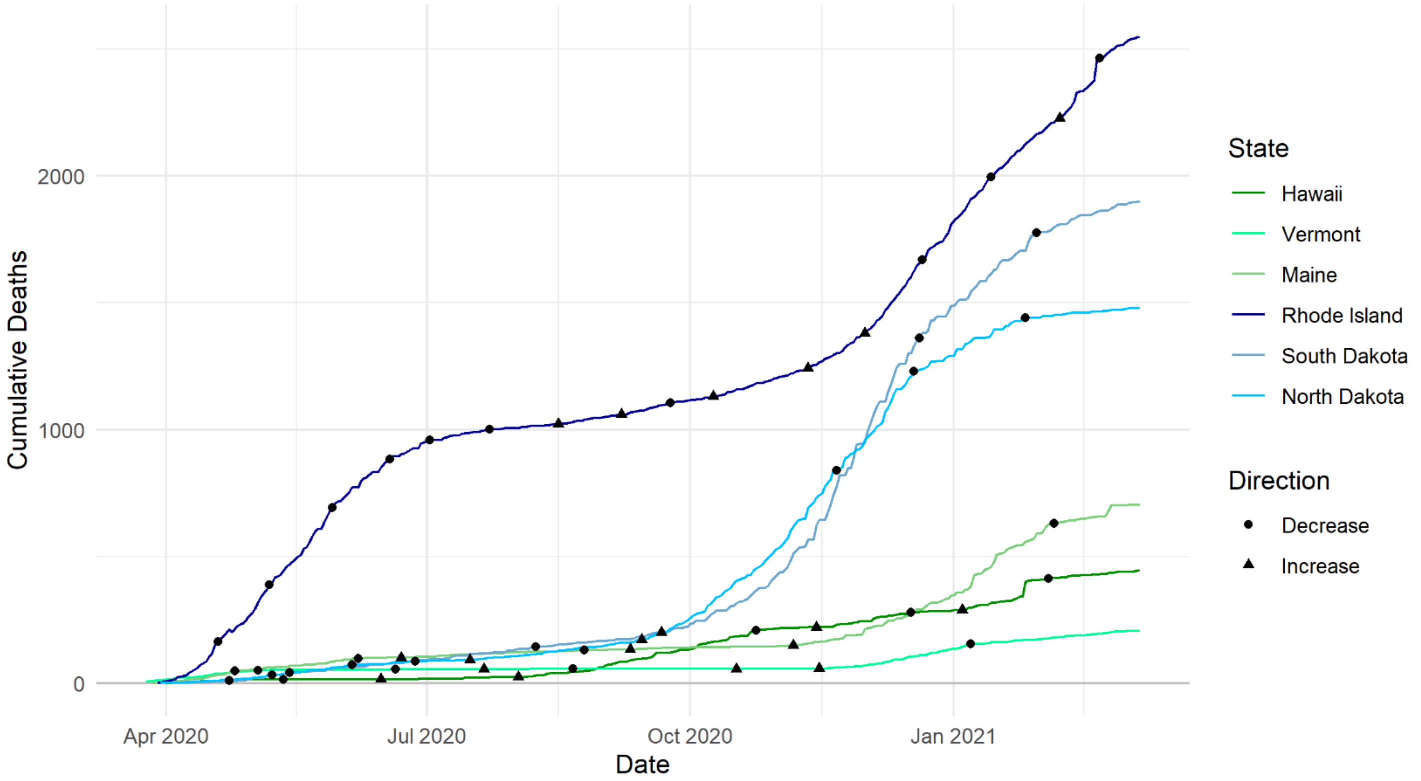

Hawaii, Vermont, and Maine ranked as the three best performing states by cumulative number of COVID-19 cases per capita as of March 7, 2021. Rhode Island, South Dakota, and North Dakota ranked as the three worst performing states over the same time period. Breakpoints, dates at which the linear segments of COVID-19 death velocities showed substantial change in their rate, are plotted over the liner plot of deaths for each respective state.

**Figure S7. Derivative of the logarithm of cumulative deaths and the breakpoints identified for the top 3 lowest and highest U.S. states by COVID-19 cases per capita**

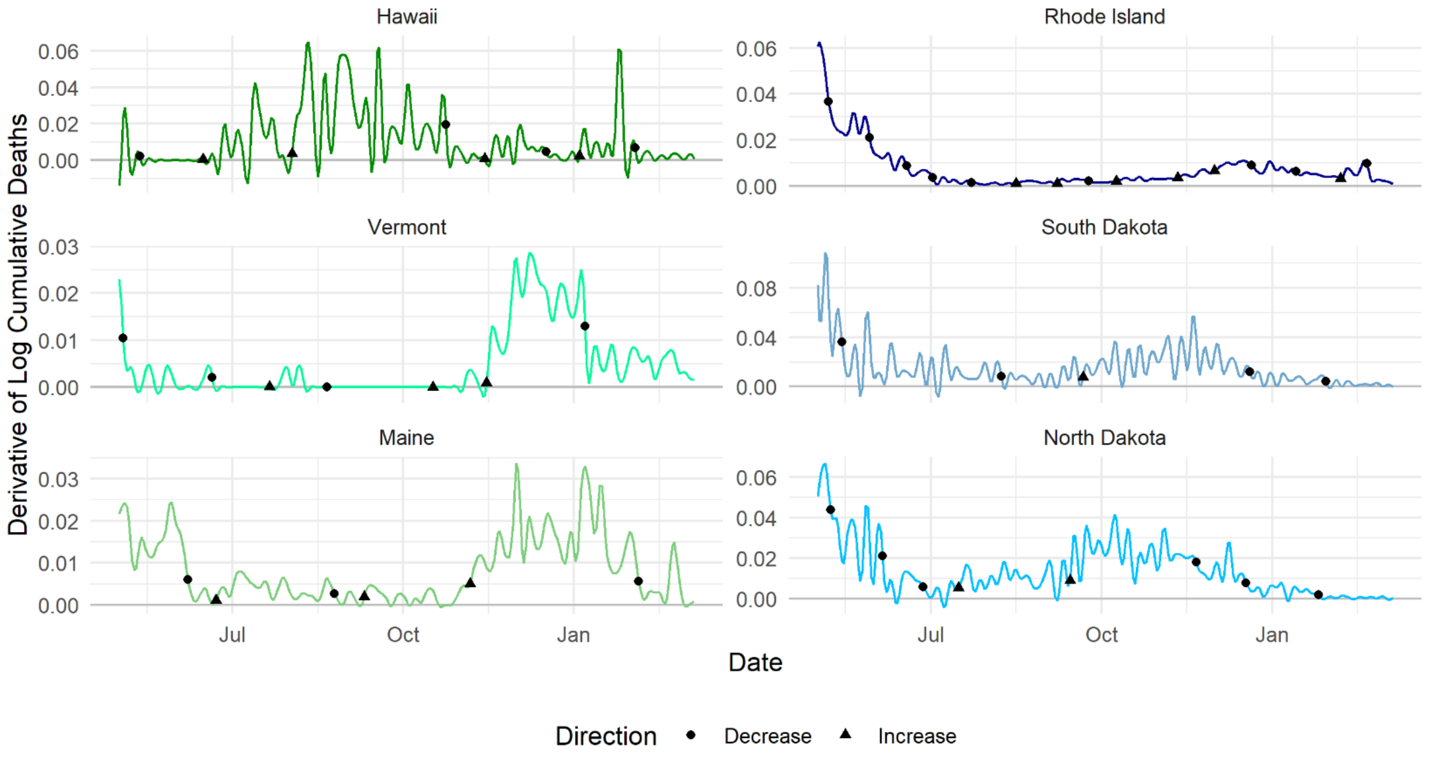

Derivative of the logarithm of cumulative deaths most closely represents case velocities graphed. X=0 for graphs were set as May 1st, 2020 rather than date of first cumulative death due to high initial velocities, pictorially obscuring clinically informative later breakpoints. Hawaii, Vermont, and Maine ranked as the three best performing states by cumulative number of COVID-19 cases per capita as of March 7, 2021. Rhode Island, South Dakota, and North Dakota ranked as the three worst performing states over the same time period. Breakpoints, dates at which the linear segments of COVID-19 death velocities showed substantial change in their rate, are plotted over the liner plot of deaths for each respective state.

**Figure S8. Logarithm of the derivative of the logarithm of cumulative deaths and the breakpoints identified for the top 3 lowest and highest U.S. states by COVID-19 cases per capita**

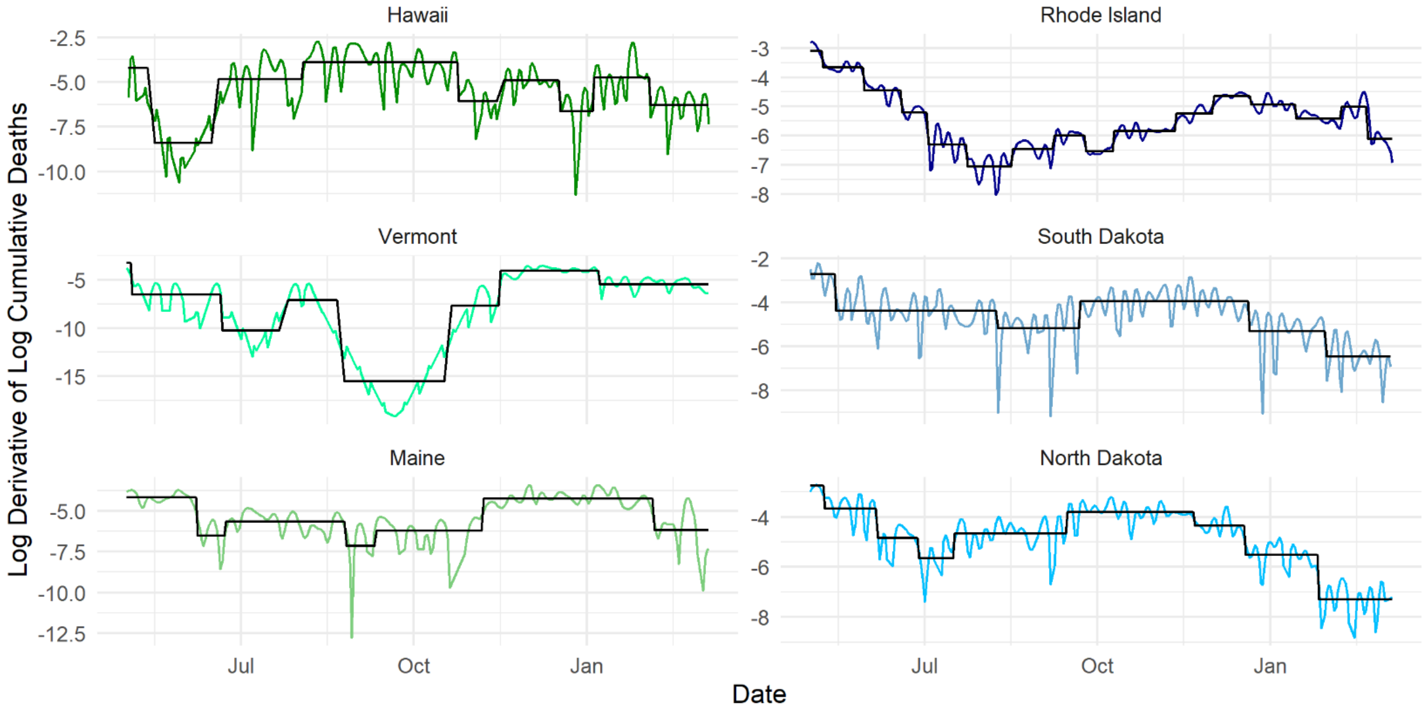

Logarithm of the derivative of the logarithm of cumulative deaths most closely represents curation of data implemented to models. X=0 for graphs were set as May 1st, 2020 rather than date of first cumulative death due to high initial velocities, pictorially obscuring clinically informative later breakpoints. Hawaii, Vermont, and Maine ranked as the three best performing states by cumulative number of COVID-19 cases per capita as of March 7, 2021. Rhode Island, South Dakota, and North Dakota ranked as the three worst performing states over the same time period. Breakpoints, dates at which the linear segments of COVID-19 death velocities showed substantial change in their rate, are plotted over the liner plot of deaths for each respective state.
